# Single dose of BNT162b2 mRNA vaccine against SARS-CoV2 induces neutralizing antibody and polyfunctional T-cell responses in patients with CML

**DOI:** 10.1101/2021.04.15.21255482

**Authors:** Patrick Harrington, Katie J. Doores, Deepti Radia, Amy O’Reilly, Ho Pui Jeff Lam, Jeffrey Seow, Carl Graham, Thomas Lechmere, Donal McLornan, Richard Dillon, Yogita Shanmugharaj, Andreas Espehana, Claire Woodley, Jamie Saunders, Natalia Curto-Garcia, Jennifer O’Sullivan, Shahram Kordasti, Michael H. Malim, Claire Harrison, Hugues de Lavallade

## Abstract

Patients receiving targeted cancer treatments such as tyrosine kinase inhibitors (TKIs) have been classified in the clinically extremely vulnerable group to develop severe acute respiratory syndrome coronavirus 2 (SARS-CoV-2), including patients with Chronic Myeloid Leukaemia (CML) taking TKIs. In addition, concerns that immunocompromised individuals with solid and haematological malignancies may not mount an adequate immune response to a single dose of SARS-CoV-2 BNT162b2 (Pfizer-BioNTech) vaccine have been raised.

We evaluated humoral and cellular immune responses after a first injection of BNT162b2 vaccine in 16 CML patients. Seroconversion and cellular immune response prior and after vaccination were assessed.

By day 21 post-vaccination, anti-Spike IgG were detected in 14/16 (87.5%) of CML patients and all developed a neutralizing antibody response (ID_50_>50), including medium (ID_50_ of 200-500) or high (501-2000) neutralising antibodies titres in 9/16 (56.25%) patients. T cell response was seen in 14/15 (93.3%) evaluable patients, with polyfunctional responses seen in 12/15 (80%) patients (polyfunctional CD4+ response 9/15, polyfunctional CD8+ T cell response 9/15).

These data demonstrate the immunogenicity of a single dose of SARS-CoV-2 BNT162b2 vaccine in most CML patients with both neutralizing antibodies and polyfunctional T-cell responses seen, in contrast to patients with solid tumour or lymphoid haematological malignancies.

**Funding:** King’s Together Rapid COVID-19 Call awards to MHM, KJD; A Huo Family Foundation Award to MHM, KJD; Chronic Disease Research Foundation award CDRF-22/2020 to KJD, MHM; Wellcome Trust Investigator Award 106223/Z/14/Z to MHM; CG was supported by the MRC-KCL Doctoral Training Partnership in Biomedical Sciences (MR/N013700/1); Fondation Dormeur, Vaduz for funding equipment to KJD

## Introduction

Severe acute respiratory syndrome coronavirus 2 (SARS-CoV-2), a novel beta coronavirus, has led to unprecedented healthcare challenges on a global scale. Development of antiviral immunity is key to reducing spread of infection and gaining pandemic control. Impressive collaborative efforts have led to the rapid development of multiple efficacious vaccines against SARS-CoV-2. BNT162b2 is a nucleoside-modified mRNA that encodes a full-length Spike that is stabilised in the prefusion conformation of the SARS-CoV-2 Spike protein, a key target of neutralising antibodies. However, concerns that immunocompromised individuals may not mount an adequate immune response following a single dose of vaccination have been raised, with early reports describing a reduced response in a heterogenous group of patients with solid and haematological malignancies^1^. Patients with CML have been shown to have impaired innate and adaptive immunity -although most pronounced at diagnosis-leading to the decision of the Department of Health and Social Care (DHSC) to classify in the clinically extremely vulnerable groups “people having other targeted cancer treatments that can affect the immune system, such as protein kinase inhibitors”. Immune response to vaccination may indeed be attenuated by tyrosine kinase inhibitors (TKI), particularly those with greater ‘off target’ kinase inhibition, which we have previously shown to inhibit B cell function and antibody response in vivo^2^. Herein we report an analysis of the T cell and antibody responses following a single dose of BNT162b2 in patients with CML on TKI therapy.

## Methods

### Study design and vaccine

From 17^th^ December 2020 until 17^th^ February 2021, patients with a known diagnosis of myeloid haematological malignancies presenting at Guy’s & St Thomas’ NHS Foundation Trust and eligible to receive a 30μg injection of SARS-COV-2 mRNA BNT162b2 vaccine were approached for informed consent. Over that period, 16 adult patients with a diagnosis of chronic Phase CML were vaccinated in compliance with UK DHSC guidelines and recruited. All patients gave informed consent, and the study protocol was approved by the regional research and ethics review board (IRAS ID: 285396; REC ID: 20/WM/0187). Peripheral blood mononuclear cells (PBMCs) and plasma were isolated pre vaccination and at a median of 21 days (IQR 21-27.25) after their first injection of 30μg BNT162b2 in 16 patients.

### Safety assessments

We solicited reports of local (pain, tenderness, redness, induration and ecchymosis) and systemic (fever, headache, malaise, myalgia, chills and nausea) adverse events by 2-weekly phone calls performed by trained medical students and physician assistant. All local and systemic adverse events reported in response to solicitation within 7 days after administration of the vaccine were considered to be related to the vaccine.

### Intracellular cytokine flow cytometry assay

T cell functionality was assessed using intracellular cytokine staining after incubation with SARS-CoV-2 specific peptides covering the immunogenic domains of the Spike (S) protein (Miltenyi Biotech). Briefly, cells were thawed, then rested for 18 hours at 37°C, 5% CO2. Specific peptides (0.25□µg/ml) and anti-CD28 (BD bioscience) were added for 3 hours, followed by Brefeldin-A (BFA) for an additional 3 hours. Unstimulated cells were utilised as negative controls and PMA and Ionomycin (Miltenyi Biotech) was added separately as a positive control. Cells were stained with a viability dye, stained with antibodies directed against surface markers, and fixed and permeabilised (BD CytoFix/Cytoperm) prior to staining with antibodies directed against intracellular cytokines. Directly conjugated monoclonal antibodies with the following specificities were used; CD3 BUV395 (clone SK37), CD4 PE (clone M-T477), CD45RO BV711, TNFα (clone MAB11), IFNγ APC (clone B27), IL-2 PE (clone MQ1-17H12). Live dead staining was performed using Zombie NIR amine reactive fluorescent dye (Biolegend). Gating on the lymphocyte population, single cells, live cells, CD3+ cells, CD4+ cells and CD4- (CD8+) was performed. T cell analysis was performed on a BD Fortessa cytometer and results processed using Flowjo version 10.5. Statistical analysis was performed using Prism, version 8.

### ELISA protocol

ELISAs were conducted as previously described^1^. All plasma samples were heat-inactivated at 56□°C for 30□min before use. High-binding ELISA plates (Corning, 3690) were coated with antigen (Nuclear (N) protein or the S glycoprotein at 3µg□ml^-1^ (25□µl per well) in PBS, either overnight at 4□°C or for 2□h at 37□°C. Wells were washed with PBS-T (PBS with 0.05% Tween-20) and then blocked with 100□µl of 5% milk in PBS-T for 1□h at room temperature. The wells were emptied and serial dilutions of plasma (starting at 1:25, 6-fold dilution) were added and incubated for 2□h at room temperature. Control reagents included CR3009 (2□µg□ml^-1^)(N-specific monoclonal antibody), CR3022 (0.2□µg□ml^-1^)(S-specific monoclonal antibody), negative control plasma (1:25 dilution), positive control plasma (1:50), and blank wells. Wells were washed with PBS-T. Secondary antibody was added and incubated for 1h at room temperature. IgG was detected using goat-anti-human-Fc-AP (alkaline phosphatase) (1:1,000) (Jackson, catalogue no. 109-055-098) and wells were washed with PBS-T and AP substrate (Sigma) was added and plates read at 405□nm. EC50 values were calculated in GraphPad Prism. Where an EC50 was not reached at 1:25, a plasma was considered seropositive if the OD at 405nm was 4-fold above background and a value of 25 was assigned.

### Neutralization assay with SARS-CoV-2

HIV-1 (human immunodeficiency virus type-1) based virus particles, pseudotyped with SARS-CoV-2 Wuhan Spike were prepared in HEK-293T/17 cells and neutralization assays were conducted as previously described^3^. Serial dilutions of plasma samples (heat inactivated at 56□°C for 30□min) were prepared in DMEM complete media (10% foetal bovine serum -FBS, 1% Pen/Strep (100 IU/mL penicillin and 100 mg/mL streptomycin) and incubated with pseudotyped virus for 1□h at 37□°C in 96-well plates. Next, HeLa cells stably expressing the ACE2 receptor (provided by Dr James Voss, Scripps Research, La Jolla, CA) were added (12,500 cells/50µL per well) and the plates were left for 72 hours. Infection level was assessed in lysed cells with the Bright-Glo luciferase kit (Promega), using a Victor™ X3 multilabel reader (Perkin Elmer). Measurements were performed in duplicate and the duplicates used to calculate the serum dilution that inhibits 50% infection (ID_50_) using GraphPad Prism.

## Results

### Patients’ characteristics and toxicity profile of SARS-CoV-2 BNT162b2

Analysis was performed in 16 chronic phase CML patients, with clinical characteristics summarised in Table 1. The vaccine was safe and tolerable in this patient group with localised inflammation reported in 56.3% (9) and a transient flu like illness reported in 23.5% (4) of patients (Table 2). None of the 16 patients reported SARS-COV-2 infection prior to enrolment or after inclusion into the study.

**Table 1:**
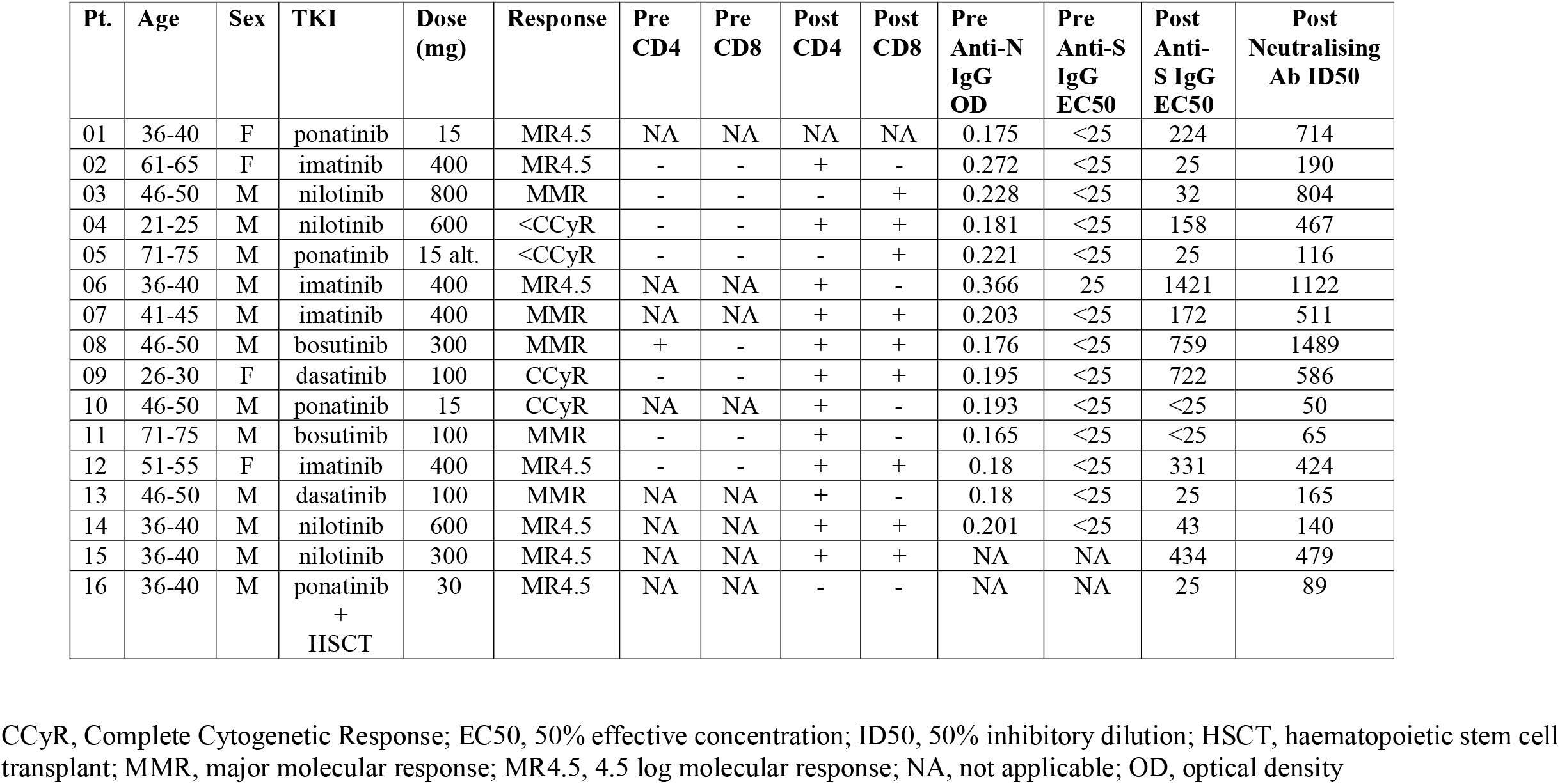
Patient’s baseline characteristics and response to first dose of vaccination.

**Table 2:** Injection-site and systemic adverse effects after receiving the vaccine among patients.

| **Pt** | **Adverse event** |  |
| --- | --- | --- |
|  | **Local** | **Systemic** |
| 01 | None | None |
| 02 | None | Flu like illness |
| 03 | Localised inflammation | None |
| 04 | Localised inflammation | Fatigue |
| 05 | Localised inflammation | None |
| 06 | Localised inflammation | None |
| 07 | None | Flu like illness<br>Headache |
| 08 | Localised inflammation | Flu like illness |
| 09 | None | Fatigue |
| 10 | None | Flu like illness |
| 11 | Localised inflammation | None |
| 12 | None | None |
| 13 | None | None |
| 14 | Localised inflammation | None |
| 15 | None | Flu like illness |
| 16 | Localised inflammation | Headache |

### Seroconversion rates to BNT162b2 and neutralising antibody titres after a first injection

Antibody testing was performed in all 16 CML patients. The presence of IgG to Spike and plasma neutralising activity was measured 3 weeks after first the vaccine injection (median 21 days, IQR 21-27.25). A positive anti-S IgG ELISA response was seen in 87.5% (14/16) of patients analysed after a first injection, with only two patients showing undetectable IgG to Spike at 1:25 dilution (Table 1 and Figure 1a).

**Figure 1.**
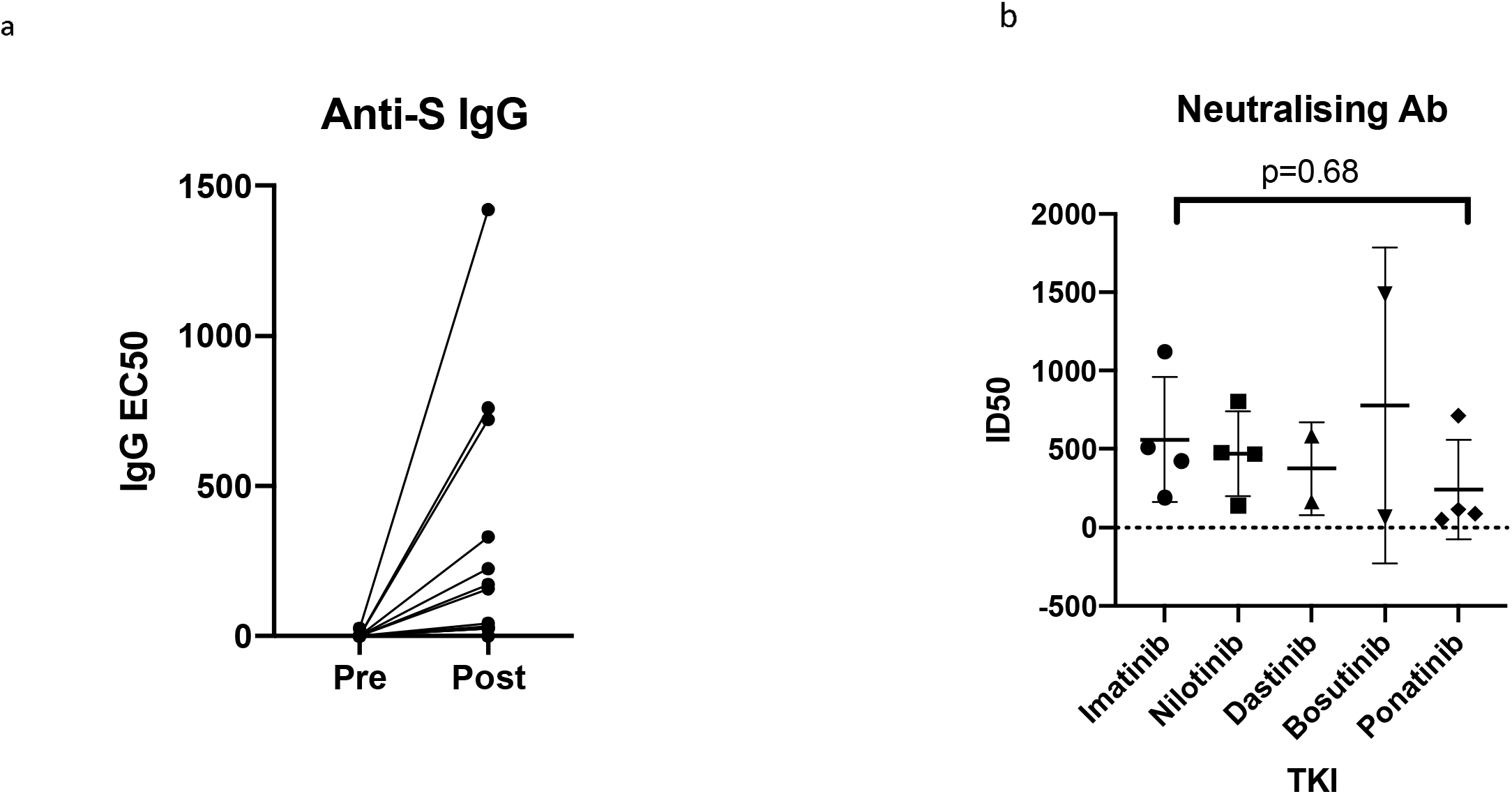
a. Anti-S IgG EC50 pre and post vaccination. b. Neutralising antibody ID50 within different TKI groups.

The median EC_50_ value for anti-S IgG in all patients from ELISA testing was 100.5 (IQR 25-408.3). Amongst different TKIs median EC_50_ values were 251.5 for imatinib, 100.5 for nilotinib, 373.5 for dasatinib, 379.5 for bosutinib and 25 for ponatinib. No patients had evidence of previous infection as determined by >4-fold increase in optical density of IgG against the Nucleocapsid (N) and S protein in the pre-vaccine sample^4^.

Neutralising antibody analysis was performed in all post-vaccine samples using an HIV-1 based virus particles, pseudotyped with SARS-CoV-2 Wuhan Spike, with positive responses seen in all patients and a median ID_50_ of 445.5 (IQR 122-682). Seven of 16 (43.8%) patients showed low (50-200), 3/16 (18.8%) medium (201-500) and 6/16 (37.5%) high (501-2000) neutralizing titres^3^.

Amongst different TKIs median ID_50_ was 467.5 for imatinib, 473 for nilotinib, 375.5 for dasatinib, 777 for bosutinib and 102.5 for ponatinib with no statistical difference seen between TKIs (Figure 1b, p=0.68). Moreover, analysis was performed after both first and second doses in one patient (Patient 01), with an increase in IgG EC50 from 224 to 4810 and neutralising antibody ID50 increase from 714 to 2037, between first and second doses.

### T-cell response to first injection of BNT162b2

The induction of virus-specific T-cell responses by BNT162b2 vaccination was assessed directly ex vivo by flow cytometric enumeration of antigen-specific CD8+ and CD4+ T lymphocytes using an intracellular cytokine assay for IFNγ, TNFα and IL2. Prior to vaccination T cell analysis was performed in 8 patients, with only one patient showing evidence of a monofunctional CD4+ T cell response to the S protein with expression of IFN gamma (Table 1).

After a first injection of BNT162b2, T cell analysis was performed in 15 patients. A response was considered to be positive if there was a 3-fold increase in any pro-inflammatory cytokine from baseline expression, and above a threshold of 0.01. A memory T cell response was seen in 14 out of 15 evaluable patients (93.3%), with the only patient not showing a T-cell response being post allogeneic haematopoietic stem cell transplantation (HSCT) and taking ponatinib (Table 1 and Figure 2). A SARS-CoV-2 specific CD4+ T cell response was seen in 80% (12/15) and a SARS-CoV-2 specific CD8+ T cell response was seen in 60% (9/15, Table 1 and Figure 2). A polyfunctional cytokine response in either CD4 or CD8+ T cells was seen in 80% (12/15) of patients, with a polyfunctional CD4+ response in 60% (9/15) and a polyfunctional CD8+ T cell response in 40% (9/15) of patients (Figure 2).

**Figure 2.**
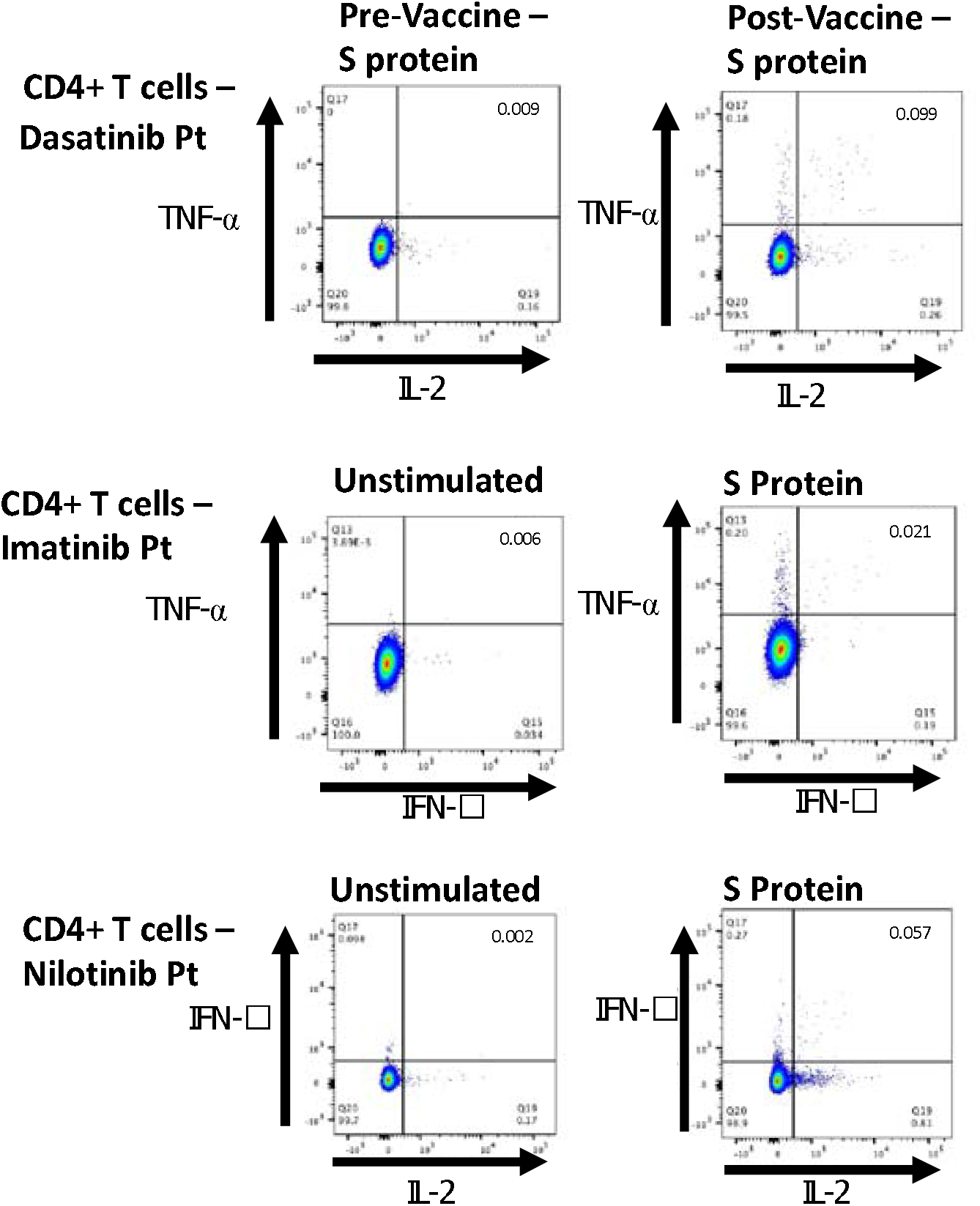
Top - Dual expression of TNF alpha and IL-2 in CD4+ T cells in dasatinib treated patient, left pre-vaccine and right post-vaccine. Middle - Dual expression of TNF alpha and IFN gamma in CD4+ T cells in imatinib treated patient, left - unstimulated cells and right - cells exposed to S protein. Bottom - Dual expression of IFN gamma and IL-2 in CD4+ T cells in nilotinib treated patient, left - unstimulated cells and right - cells exposed to S protein.

The median increase in expression of TNFα in CD4+ cells compared with the baseline unstimulated control was 0.071 (IQR 0.039-0.25) and in CD8+ cells 0.032 (IQR 0.004-0.6). IFNγ was 0.027 (IQR 0-0.11) in CD4+ and 0.091 (IQR 0.007-1.18) in CD8+ cells whilst median IL-2 expression was 0.05 (IQR 0.05-0.11) in CD4+ cells and 0.01 (IQR 0.056-0.09) in CD8+ cells. With regards to polyfunctional responses, the median increase in TNFα+/IFNγ+ cells was 0.004 (IQR 0.002-0.014) in CD4+ cells and 0.007 (IQR 0-0.039) in CD8+ cells, with a median increase in expression of TNFα+/IL-2+ of 0.05 (IQR 0.05-0.11) in CD4+ cells and 0.002 (IQR 0-0.004) in CD8+ cells. Greater than 90% of reactive cells expressing IFNγ or TNFα co-expressed CD45RO, consistent with a memory T cell phenotype. Interestingly patients taking nilotinib had a significantly higher mean increase in TNFα expression in CD4+ cells, when compared with patients taking other TKIs (0.83 vs 0.096, p=0.015, Figure 3a), and also of dual TNFα+/IFNγ+ (0.098 vs 0.009, p=0.028, Figure 3b). No other significant differences between TKIs were identified.

**Figure 3.**
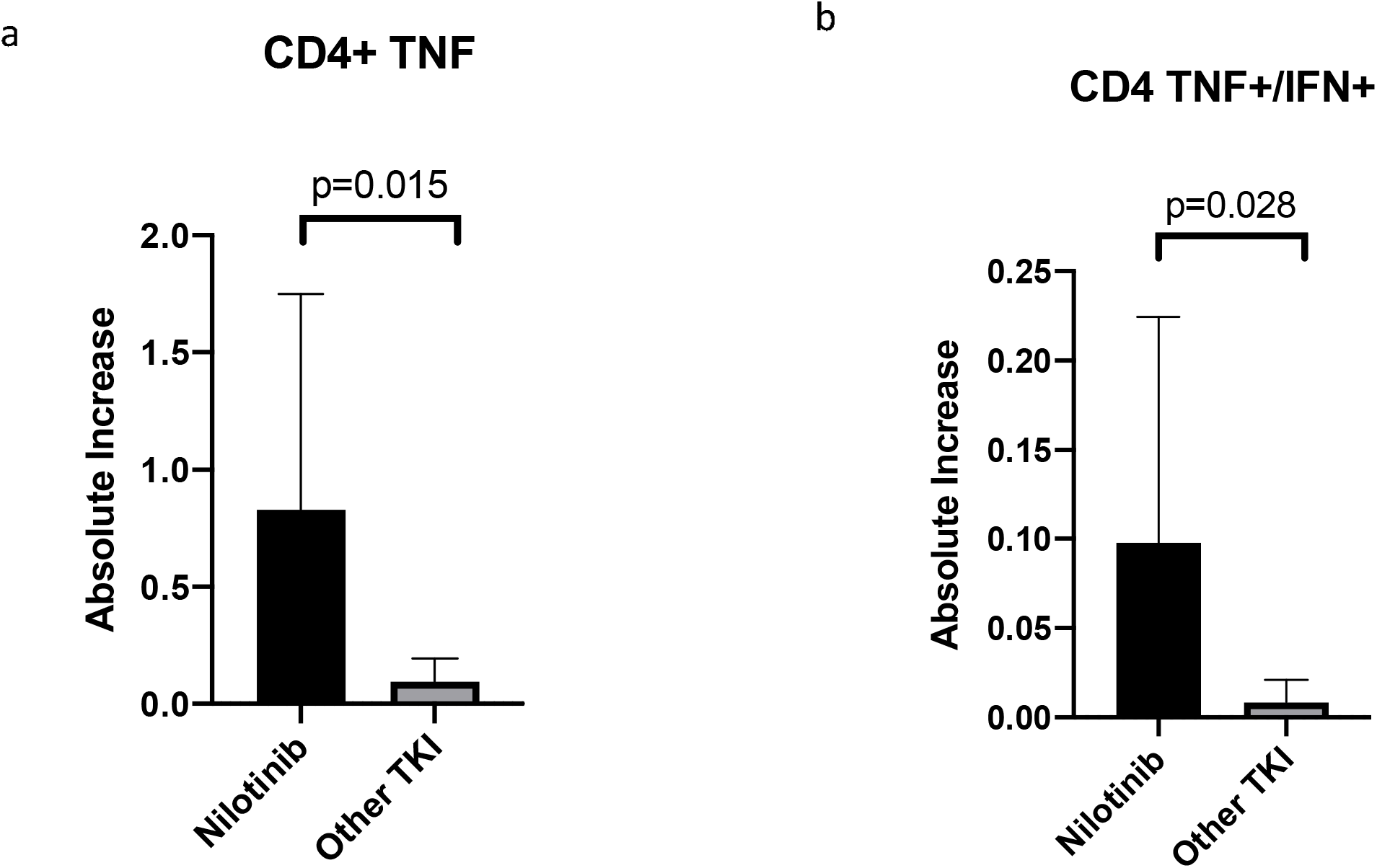
a. Mean increase in CD4+ T cell TNF alpha expression in nilotinib treated patients compared with other TKIs. b. Mean increase in CD4+ T cell dual TNF alpha and IFN gamma expression in nilotinib treated patients compared with other TKIs.

## Discussion

Large international randomised placebo-controlled studies have demonstrated the efficacy of the BNT162b2 vaccine, with a two-dose regime resulting in a 95% reduction in cases in the general population^5^. Evidence of a protective immune response to a single dose is shown by a reduction in cases of 52% reported in the interim between first and second doses in those receiving BNT162b2^5^. Sahin and colleagues reported a detailed analysis of the immune response to BNT162b2 in healthy controls, with a Th1 skewed CD4+ T cell response in 95.2% and a CD8+ T cell response in 76.2%, as determined by Elispot testing^6^. They also described antibody responses to vaccination that were significantly greater than that observed in a group of patients who had recovered from SARS-CoV-2 infection^6^.

T cell immunity may prove to be particularly important for protection from SARS-CoV-2, with the evidence from the SARS-CoV-1 epidemic showing that antibody responses were relatively short-lived, following infection, whilst memory T cell responses were significantly more durable.^7^ Our group has recently reported a significant decline in neutralising antibody levels in the 3 months after infection with SARS-CoV-2.^3^ We have also demonstrated that robust T cell responses can be observed as long as 6 months after infection with SARS-COV-2 in a small cohort of patients with chronic myeloproliferative neoplasms including CML.^8^ Furthermore, a number of additional studies have elegantly demonstrated the T cell response to SARS-CoV-2 in the general population.^9-13^

However, a recently submitted report describes a significantly reduced immune response in a heterogenous group of patients with solid and haematological malignancies,^1^ bringing concerns over the ability of clinically vulnerable patients with haematological malignancies to mount a protective response following a single dose of SARS-CoV-2 BNT162b2 vaccination.

Among patients with chronic myeloid malignancies, TKI therapy is recognised to cause variable immune deficiency, with TKIs that also inhibit Src family kinases causing significant T cell dysfunction due to the pivotal role that these kinases play in signalling downstream from the T cell receptor^14^. We have previously shown that B cell function is also impaired by TKI therapy through off target inhibition of Btk, a kinase that is important for normal B cell signalling, leading to reduced efficacy of the seasonal influenza vaccination.^2^ However, during the H1N1 vaccination campaign we demonstrated that the seroprotection rate to a first injection of 2009 H1N1 vaccine (Pandemrix GSK, UK) was not significantly different between patients with chronic phase CML and healthy controls, unlike other haematological conditions such as B-cell malignancies or recipients of HSCT.^15^

In line with our previous experience in H1N1 and seasonal influenza vaccination, our data show encouraging results with 87.5% seroconversion to a first SARS-CoV-2 BNT162b2 injection in CML patients, contrasting with what has been described in the first report of vaccination in cancer patients^1^. In addition, nearly all CML patients in our analysis have demonstrated both monofunctional and polyfunctional T cell cytokine responses to SARS-CoV-2, a response that is similar to healthy controls tested in previous trials of BNT162b2 vaccination^6^. Overall, both seroconversion and T-cell response to a single injection demonstrate the immunogenicity of SARS-CoV-2 BNT162b2 in CML patients, with neutralising antibodies responses seen in all patients. To our knowledge this is the first report of humoral and T-cell response to vaccination against SARS-CoV-2 in patients with CML taking TKIs. These data are particularly important in the context of the decision from the DHSC to increase the interval between a first and a second vaccine injection and demonstrate that CML patients taking TKIs receive significant protection from a first injection of SARS-CoV-2 BNT162b2.

Finally, we found that patients taking nilotinib had a significantly higher increase in TNFα and dual TNFα/IFNγ expression in SARS-CoV-2 specific CD4+ cells, when compared with patients taking other TKIs. This suggests that the highly selective ABL1 TKI nilotinib, which does not inhibit Src kinases or other kinases important to the immune response, might have minimal effect on the patients’ immune system.

Further prospective data are however required to confirm that the immune response to vaccination we have described translates into a reduction in SARS-CoV-2 infections in this patient group. In addition, longitudinal studies are required to characterise the response to the two-dose vaccination regimen since a second injection may provide a stronger and longer lasting protection. Additional studies will need to determine if the immune response will be sufficient to protect against emerging variants, as well as analyse the effect of other currently available and emerging vaccines against SARS-CoV-2.

## Data Availability

Data available within the article or its supplementary materials

## Acknowledgements

PH and HdL designed the research, performed the research, analysed the data and wrote the manuscript. KJD, JSe, CG, TL and MM perfomed the research and reviewed the manuscript. DR, HPJL, RD, CW, JSa, NCG, JOS and SK assisted with patient recruitment and reviewed the manuscript. AOR, YS and AE assisted with patient recruitment, patient interviews and reviewed the manuscript. DM and CH designed the research, assisted with patient recruitment and reviewed the manuscript.

